# Rhythmic components of COVID-19 daily cases in various countries

**DOI:** 10.1101/2020.07.23.20161240

**Authors:** V.A. Drozdov, T.A. Zenchenko

## Abstract

Not only does COVID-19 pandemic encourage scientists to look for remedies and treatment schemes, but also identify the drivers of pathogenicity and spread of the virus.

The scope of this research consisted in identifying recurrence patterns and comparing the number of daily cases between various countries. Data for countries where at least 500 daily cases were recorded at least once (17 in Europe, 3 in North America, 7 in South America, 3 in Central America, 17 in Asia and 3 in Africa).

According to our evaluation, the dynamics recorded for 25 countries includes a 7-day statistically significant component. This statistically significant weekly component has been identified in 76% of the countries examined in Europe, 66% in North America, 71% in South America, and 18% in Asia. The range of this rhythmic component is low at the growth stage and increases at the stabilization and decrease stages.

The weekly phases feature shifting peaks depending on the country. In some cases, the phases shift, i.e. they are not limited strictly to certain days of the week. Due to range and phase variation, its explanation cannot be limited to strictly medical and social factors. In some cases, national incidence dynamics includes 3, 4, 6, 8 and 10-day periods.

Understanding the factors of recurrence patterns in COVID-19 incidence dynamics may help in the pandemic response.

## INTRODUCTION

Studying COVID-10 behaviour in various countries and identifying its rate and spread drivers are among the most relevant tasks. The solution of these challenges is necessary both for proper prediction at the practical level and for identifying disease origin and spread patterns at the fundamental level.

Pandemic response decision-making is frequently related to predictions produced using various computing models. Such models are based on specific input. The authors of some recent publications have made attempts to identify COVID-19 drivers [1, 2]. For example, study [1] offers a statistical analysis of weather-related increase in daily cases. The authors conclude that disease rate is related to average temperature.

The scope of this study consisted in assessment of COVID-19 daily case increment dynamics in various countries in order to identify rhythmic components. Identification of general and specific features in disease dynamics is expected to further insight into the internal logic of the pandemic spread. This will make prediction, i.e. assessment of peaks, stabilization and case rate decrease, possible.

## MATERIALS AND METHODS

### Data sources

Daily COVID-19 case data were obtained from the web site of Center for Systems Science and Engineering (CSSE) at Johns Hopkins University (JHU) (https://systems.jhu.edu/research/public-health/ncov/) and from Google database of national agency data (“coronavirus statistics by country” search query). Data verification for each country was performed through matching data from both sources.

### Country selection criteria

The shortlisting criterion required at least a single daily case rate increment equal to or exceeding 500 at least once within the review period (01.03.2020 - 13.06.2020, 105 days), and the number of zero case days in the above databases not exceeding 3. This allowed shortlisting the countries where the statistical spread of data did not have significant impact on the daily increment rates.

Hence, 50 countries with relatively equal distribution across Europe, America and Asia were included in the sample. The full list of countries is provided in Table 1.

**Table 1.**
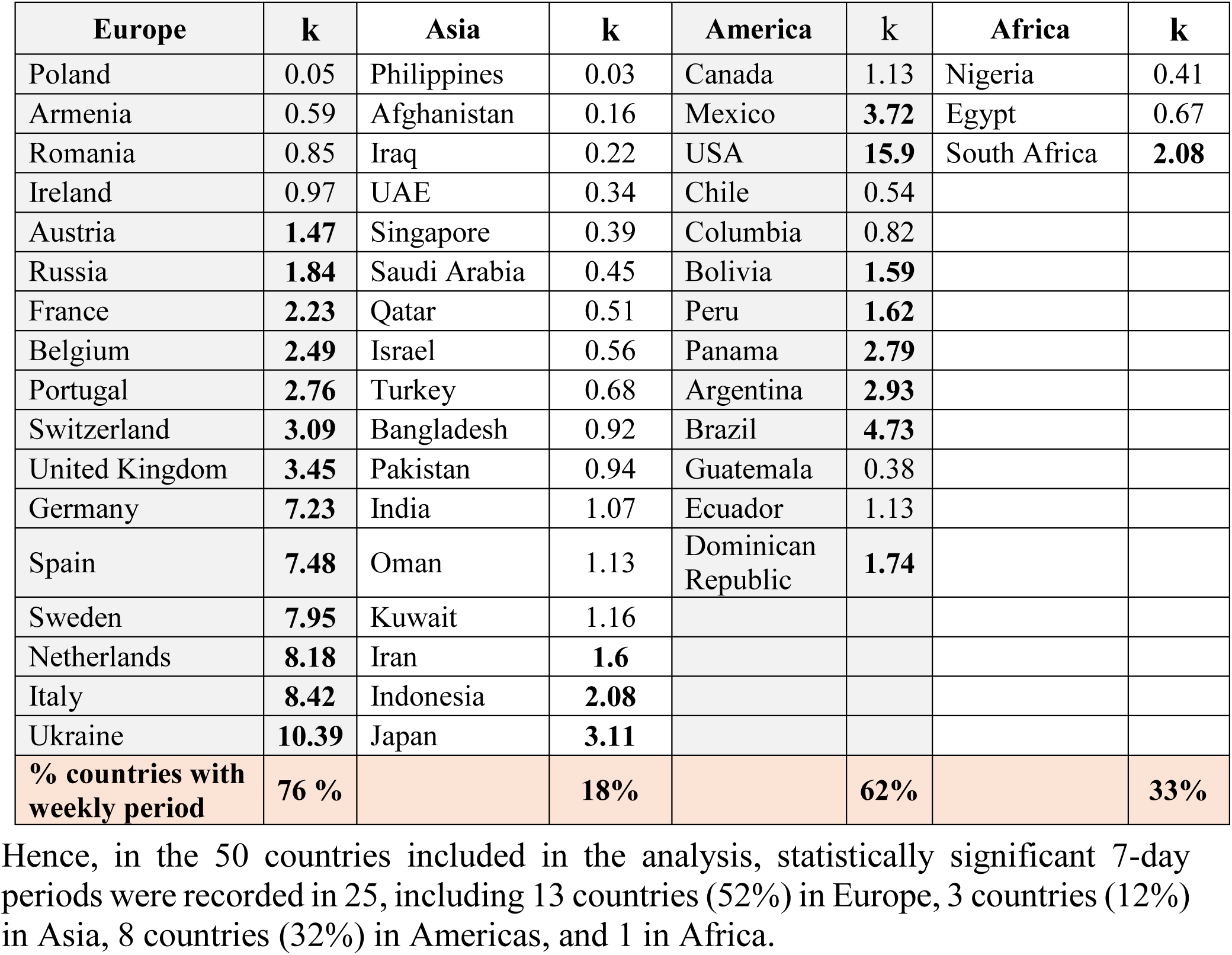

| Europe | k | Asia | k | America | k | Africa | k |
| --- | --- | --- | --- | --- | --- | --- | --- |
| Poland | 0.05 | Philippines | 0.03 | Canada | 1.13 | Nigeria | 0.41 |
| Armenia | 0.59 | Afghanistan | 0.16 | Mexico | **3.72** | Egypt | 0.67 |
| Romania | 0.85 | Iraq | 0.22 | USA | **15.9** | South Africa | **2.08** |
| Ireland | 0.97 | UAE | 0.34 | Chile | 0.54 |  |  |
| Austria | **1.47** | Singapore | 0.39 | Columbia | 0.82 |  |  |
| Russia | **1.84** | Saudi Arabia | 0.45 | Bolivia | **1.59** |  |  |
| France | **2.23** | Qatar | 0.51 | Peru | **1.62** |  |  |
| Belgium | **2.49** | Israel | 0.56 | Panama | **2.79** |  |  |
| Portugal | **2.76** | Turkey | 0.68 | Argentina | **2.93** |  |  |
| Switzerland | **3.09** | Bangladesh | 0.92 | Brazil | **4.73** |  |  |
| United Kingdom | **3.45** | Pakistan | 0.94 | Guatemala | 0.38 |  |  |
| Germany | **7.23** | India | 1.07 | Ecuador | 1.13 |  |  |
| Spain | **7.48** | Oman | 1.13 | Dominican Republic | **1.74** |  |  |
| Sweden | **7.95** | Kuwait | 1.16 |  |  |  |  |
| Netherlands | **8.18** | Iran | **1.6** |  |  |  |  |
| Italy | **8.42** | Indonesia | **2.08** |  |  |  |  |
| Ukraine | **10.39** | Japan | **3.11** |  |  |  |  |
| **% countries with weekly period** | **76 %** |  | **18%** |  | **62%** |  | **33%** |

## Method

For rhythmical component identification spectrography, wavelet transform and periodogram method were used.

For LF trend trimming the signal was pre-filtered using a band-pass filter with a Blackman-Harris window the lower and upper cutoff bandwidth set at 0.1 and 0.95 Nyquist bandwidth respectively.

For comparing results obtained using different tools, the scale parameters obtained through wavelet transform were converted to temporal characteristics similar to oscillation periods in spectrography.

In the periodogram analysis, statistical significance of the reviewed periods was assessed using Wilcoxon distribution-free test.

## RESULTS

### Global

Figure 1 shows the global dynamics of COVID-19 daily case increment and the results of the time series analysis using three approaches: periodogram (fig. 1b), wavelet transform (fig. 1c) and Fourier transform (fig. 1d).

**Figure 1.**
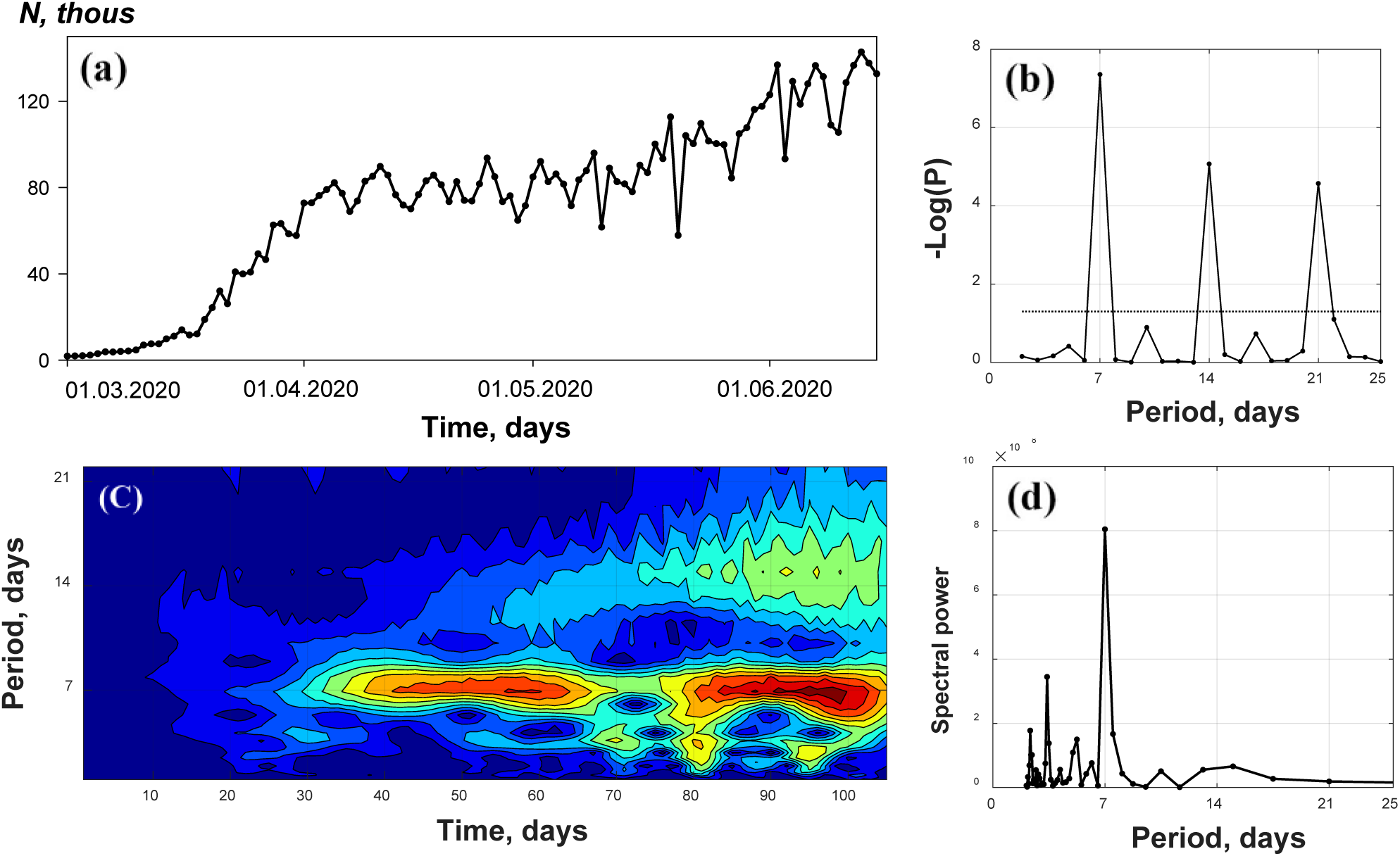
COVID-19 global daily case rate increment from 01.03.2020 through 13.06.2020. See comments in the text.

Figure 1a shows the absolute incidence values for the previous 24 hours. Four time periods featuring different growth rates are identifiable: (1) a very slow growth at the initial stage, (2) a rapid increase from 17.03 through 03.04, (3) a plateau from 03.04 through 15.05, and (4) final growth from 15.05 through the end of the period.

Figure 1b shows a periodogram with the test period plotted in days over the X-axis and negative statistical significance log taken over the Y-axis. Y-values exceeding 7 mean that a weekly period has a statistical significance of p<10^−7^, while its harmonics are 14-day (p<10^−5^) and 21-day (p<10^−4^) periods. The dotted line indicates a 5% level of statistical significance (-lg(p)=1.3).

Figure 1c shows the wavelet transform results. X-line is the day scale effective 1 March 2020, and Y-line is the test period scale, the spectral power being shown by coloured areas. Times scales of figures 1a and 1c match.

All three rhythmic component tests demonstrate the presence of a solid 7-day period. 2nd and 3rd order harmonic components are also evident in the periodogram. Wavelet and Fourier analysis also reveal periods of about 3 and 15 days. It is arguable that it is autonomous, rather than a 2nd order harmonic of the weekly period by periodicity,since the wavelet transform, contrarily to Fourier analysis, has not revealed any multiple harmonics.

A comparison of figures 1a and 1c reveals that any periodicity is absent in period 1, where the absolute values are low, with gradual spectral power increase at interval 2, and is even stronger at intervals 3 and 4, with a slight boundary decrease of the power. Similar behaviour has been recorded for the 3-day period, despite being much lower in power. At stage 4 a 15-day period arises and gradually acquires power.

As can be seen from figure 1c, the three and seven day periods have been constant for two months. Their stability in the dynamics of the global incidence time series, which is the sum of national autonomous series, is indicative of its existence and need for an advanced analysis.

### Analytical results for 50 countries

In the periodicity analysis of incidence series for 50 countries, we used the level of its statistical significance in the periodogram method with the boundary value of p<0.05 (-lg(p)>1.3, fig. 1 b). Table 1 represents country samples by continent and shows the statistical significance of the 7-day period as k=-lg(p).

One of the most likely explanations of the 7-day period could have a social nature, i.e. inconsistent reporting of new cases on weekdays versus weekends. This hypothesis was tested by plotting the phases of this period at the global level and for 15 countries with the most evident weekly period (fig. 3). Since the beginning of the review interval falls on 1 March (Sunday) and the source data reflect incidence increment reported for the *previous* 24 hours, the first two points on the X-line correspond to weekend days in figure 3 distribution patterns.

As follows from figure 2, phasal dips and peaks occur on different days of the weekly period depending on the country. The hypothesis related to 7-day period formation due to decrease in case reporting on weekends can substantiate the dynamics only for some countries, e.g. USA, Sweden, Brazil and UK. For the rest of the countries this hypothesis does not provide sufficient substantiation. For example, it does not match the global weekly periodicity with dips on Thursdays.

**Figure 2.**
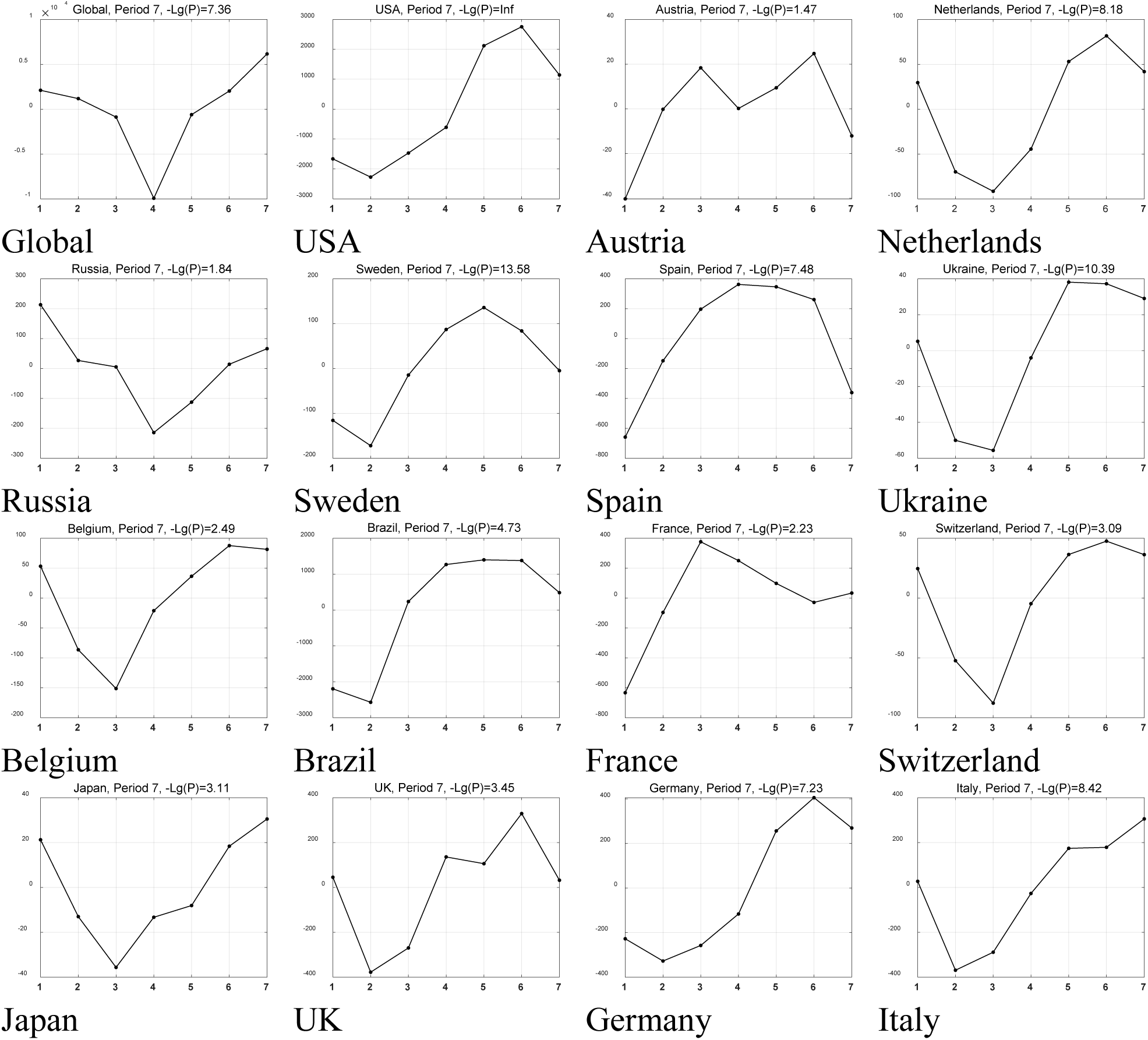
National weekly period phases

### Phase coincidence examples

Another argument against the medical and social nature of weekly periodicity consists in comparing the dynamics between various countries. Figure 3 shows the comparison between two countries with strongly manifested weekly periods, i.e. the Netherlands and Spain. This chart includes: (a) superposition of raw time series, (b) superposition of filtered time series, (c) cross correlation between them, (d) sliding window correlation by 30 points, (e) and (f) wavelet and Fourier images of the filtered series for Spain, (g) and (h) equivalent images for the Netherlands. The superposed time series are shifted against each other by 2 days in figures 3a and 3b.

As follows from figure 3, the pandemic spread phases almost coincide in these two countries. The curves in figures a and b are shifted against each other by 2 days in order to better demonstrate the similarity of the weekly phases through the shift. The existence of the shift is evidenced by cross-correlation, where the peak value of the function corresponds to −2 days (Fig. 3c). Figure 3d shows a 30 point sliding window correlation between the two filtered series. Here one can see that there is virtually no correlation between the series during the first 60 days, or that it is rather negative due to the phasal differences. However, the correlation ratio increases significantly at the end of the observation period. This occurs when the oscillations in these two series synchronize.

Hence, even in incidence time series where the weekly period is the sole and highly statistically significant one, its phase does not remain constant in some cases.

It is notable that phase periodicity does not depend on the national quarantine policy, since similar oscillation phases have been recorded for Sweden and Germany, although not special quarantine restrictions have been introduced in Sweden, by contrast with Germany. Similar findings have been recorded in the comparison of other countries differing both in terms of quarantine restrictions and socio-cultural features.

### Other periods

Despite the fact that the 7-day period has been recorded in 50% of the countries included in the sample, presence of other stable periods has been recorded in some cases. The examples of such countries are provided in figure 4.

**Figure 4.**
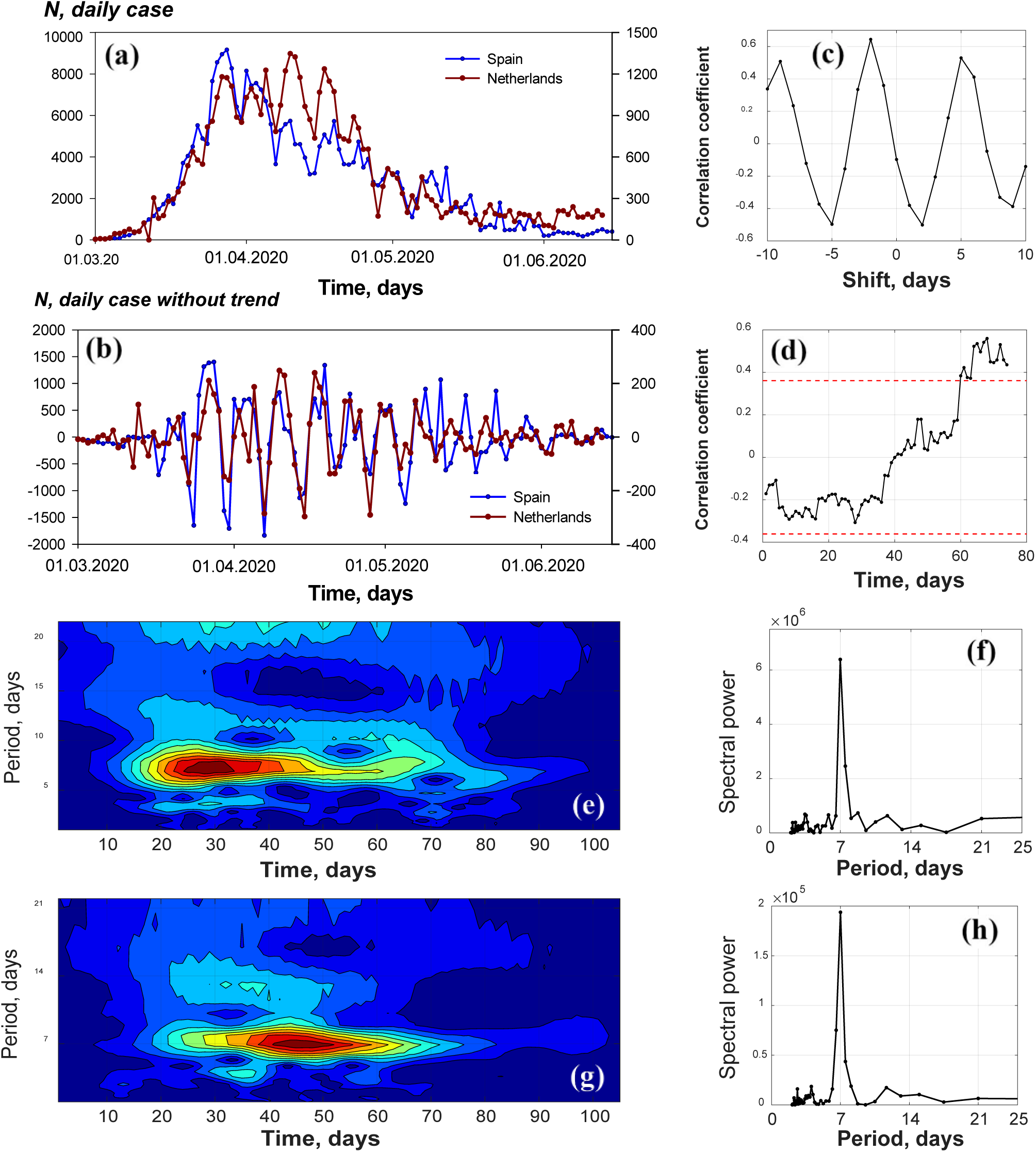
COVID-19 incidence comparison between Spain and the Netherlands. See comments in the text. The values for the Spanish series are shifted forward by 2 points in figures (a) and (b).

**Figure 5.**
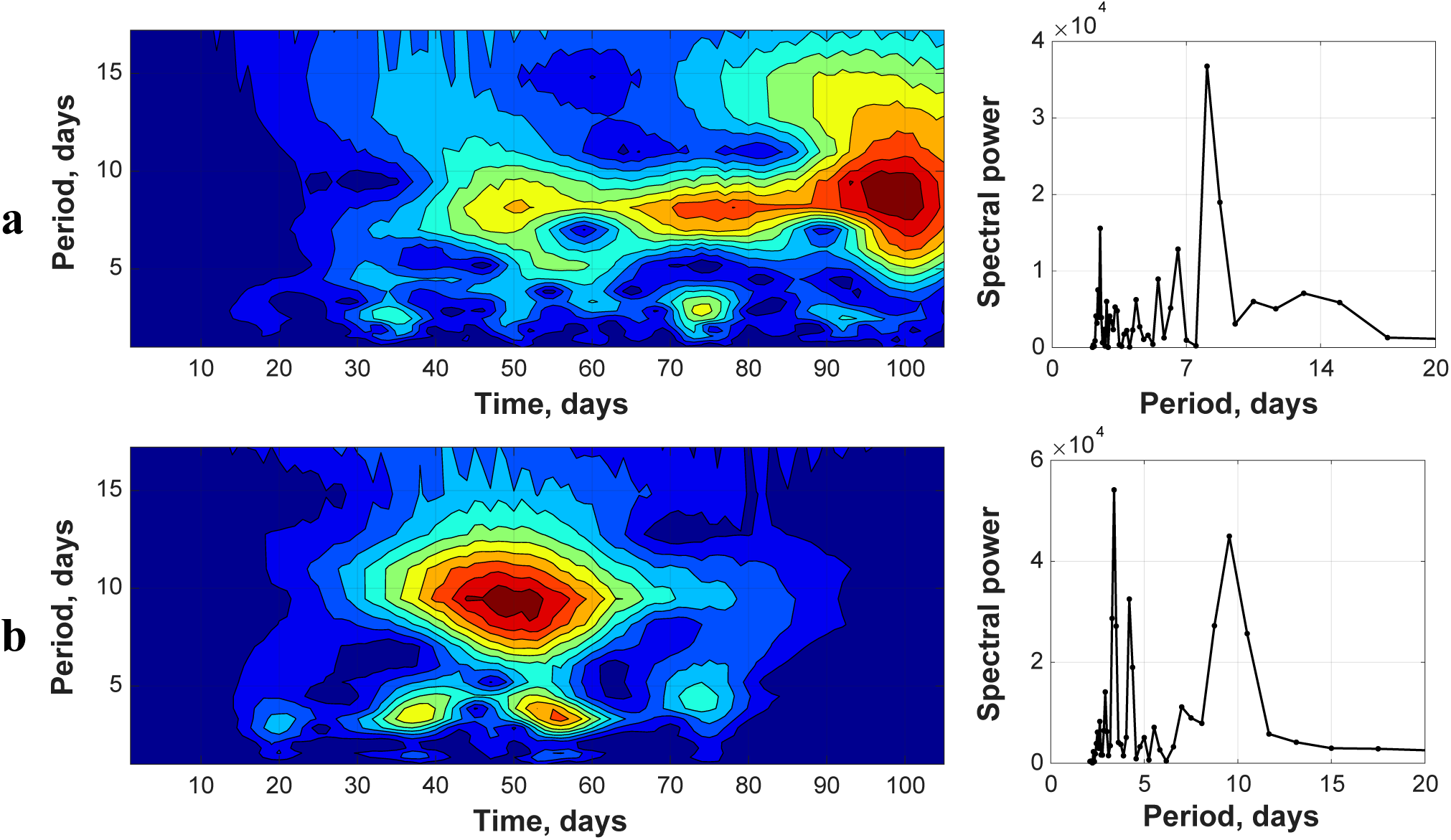
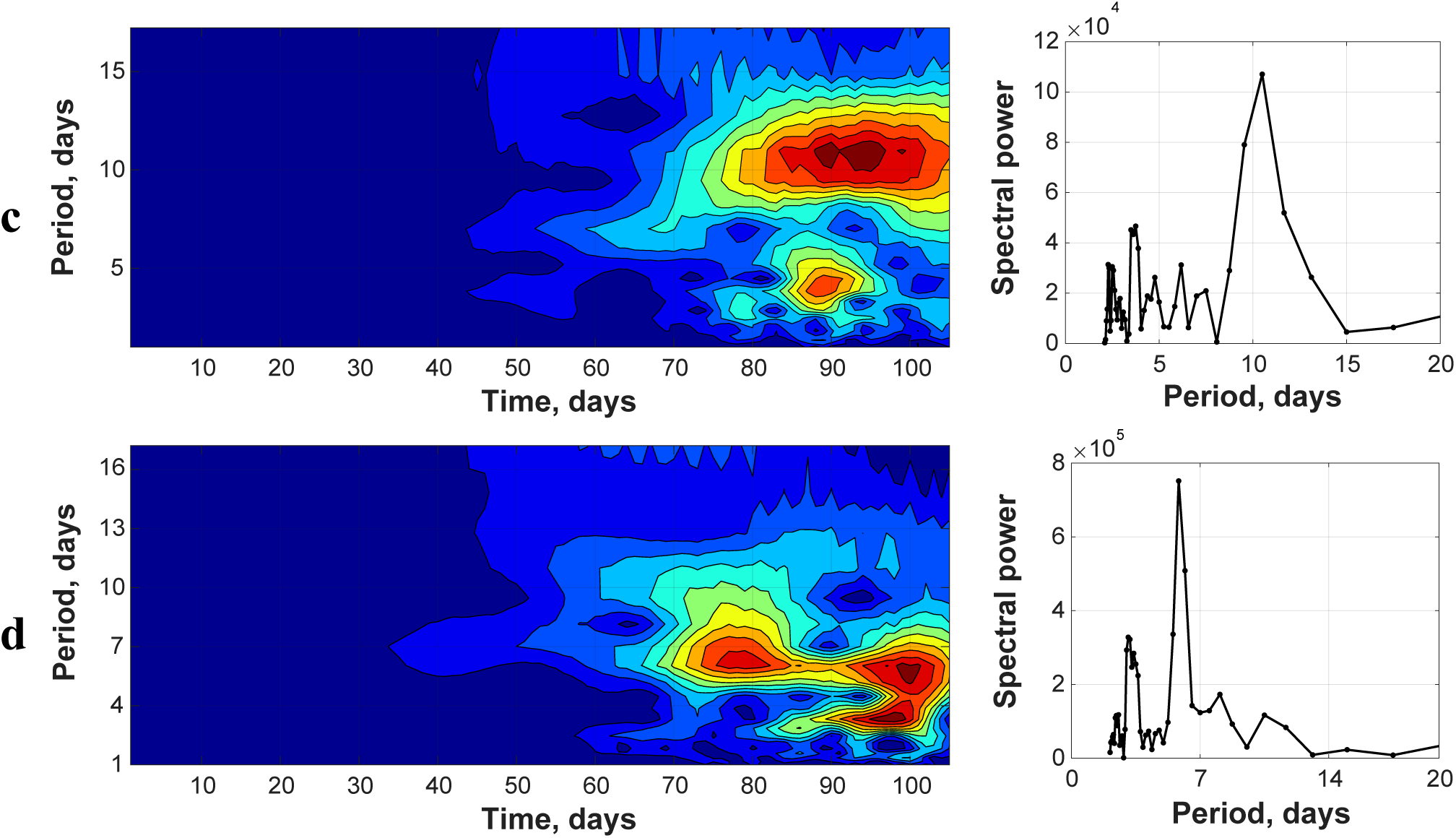
Wavelet and Fourier images of incidence time series for countries with other than weekly incidence dynamics periods. (a) - Poland, (b) - Ireland, (c) - Bangladesh, (d) - Chile.

The examples in figure 4 show an 8-day period in Poland that has been lasting for 2.5 months since the outbreak (early April), and another, less intensive, 3-day period. In Ireland, where the incidence growth period was rather short and occurred between April and early May, 10-day and 4-day periods have been recorded throughout the entire increment interval.

Chile and Bangladesh are among the countries with current rapid increase in daily incidence. Bangladesh dynamics features 10 and 4 day periods, while Chile - 6 and 3 day periods. An advanced study of the dynamics and identification of the factors related to such periodicity should become the focus of further exploration.

## DISCUSSION AND FINDINGS

A strong weekly period has been identified in 25 countries from the 50 included in the sample, such weekly period manifesting more evidently at the incidence peak and decline phases. The weekly period has been identified primarily in European countries (76% of the countries included in the sample), as well as in North America (66%) and South America (71%), yet seldom in Asia (18% of the countries included in the sample). Drawing any conclusions about Africa is difficult, since only three African counties met the sampling criteria.

The most simple and convincing explanation of the origin of the weekly period in the incidence dynamics seems to be related to varying levels of epidemiological activity and admission to hospital over weekdays versus weekends. To some extent it can be recorded for the relevant structures of all countries included in the sample. Furthermore, other social or economic factors related to irregularity of case reportingover the weekly period may exist. However, the factors of phase consistency may only partially explain the effect, while the rest of it does match this hypothesis.

1. For example, in case of inconsistent intensity of medico-social service activity, the phase of the effect should remain constant throughout the entire observation period, with the dip occurring on Saturdays and Sundays. As follows from figures 3 and 4, this is not true in many cases. Weekly period phase shifts over the observation period have been recorded in some cases.
2. The weekly period begins to manifest consistently closer to incidence peak and generally does not manifest at the initial stage. Were this caused by diagnosing irregularity, periodic existence would not be related to the pandemic phase in a given country.
3. The weekly period has been detected in the incidence dynamics of countries with various health care systems and various pandemic response strategies, including both quarantine-based and quarantine-free (see Table 1).
4. In the incidence dynamics of some countries consistent and statistically significant periods other than weekly have been recorded, i.e. 6, 8 or 10 days.

Hence, the identified periodicity patterns are not accounted for solely by the investigated medico-social factors. One could presume that some less obvious medico-biological aspects related to the viral spread underly the observed periodicity patterns. This problem deserves advanced investigation, since the findings of such investigation could help to explain the specifics of viral spread in various countries.

## Data Availability

All data referred to in the manuscript are available

